# Comparison of the reactogenicity and immunogenicity of a reduced and standard booster dose of the mRNA COVID-19 vaccine in healthy adults after two doses of inactivated vaccine

**DOI:** 10.1101/2022.03.01.22271735

**Authors:** Sitthichai Kanokudom, Suvichada Assawakosri, Nungruthai Suntronwong, Jira Chansaenroj, Chompoonut Auphimai, Pornjarim Nilyanimit, Preeyaporn Vichaiwattana, Thanunrat Thongmee, Ritthideach Yorsaeng, Thaneeya Duangchinda, Warangkana Chantima, Pattarakul Pakchotanon, Donchida Srimuan, Thaksaporn Thatsanatorn, Sirapa Klinfueng, Juthathip Mongkolsapaya, Natthinee Sudhinaraset, Nasamon Wanlapakorn, Sittisak Honsawek, Yong Poovorawan

## Abstract

The coronavirus disease 2019 (COVID-19) pandemic has been a serious healthcare problem worldwide since December 2019. The third dose of heterologous vaccine was recently approved by World Health Organization. The present study compared the reactogenicity and immunogenicity of the reduced and standard third booster dose of the BNT162b2 and mRNA-1273 vaccine in adults who previously received the two-dose CoronaVac vaccine. Results showed that headache, joint pain, and diarrhea were more frequent in the 15 μg-than the 30 μg-BNT162b2 groups, whereas joint pain and chilling were more frequent in the 100 μg-than the 50 μg-mRNA-1273 groups. No significant differences in immunogenicity were detected. These findings demonstrate that the reduced dose of the mRNA vaccines elicited antibody responses against the SARS-CoV-2 delta and omicron variants that were comparable to the standard dose. The reduced dose could be used to increase vaccine coverage in situations of limited global vaccine supply.

**Highlights:**

- The 15 μg- and 30 μg-BNT162b2, and 50 μg- and 100 μg-mRNA-1273 booster doses were compared
- Booster vaccination with the mRNA vaccine elicits high Ig and IgG anti-RBD in CoronaVac-vaccinated adults
- No differences were observed in antibody responses after the reduced or standard booster dose of the mRNA vaccine in CoronaVac-vaccinated adults
- Neutralizing antibodies against the delta and omicron variants were significantly higher after the booster dose
- Neutralizing antibody titers were lower against the omicron variant than the delta variant in all vaccinated adults

## 1. Background

A shortage of the coronavirus disease 2019 (COVID-19) vaccine has been reported in several countries [2]. Vaccine importing countries, including Thailand, are affected by vaccine shortage, resulting in delays in the vaccination schedule. A dose reduction strategy can help countries to save costs on the imported vaccine and distribute vaccine more quickly during the COVID-19 outbreak. Several countries achieved high two-dose COVID-19 vaccination coverage but have been unable to control the spread of SARS-CoV-2. The delta and omicron variants have recently spread worldwide [3], and are causing breakthrough infection in vaccinated individuals [4, 5]. A preliminary study showed that vaccination of inactivated vaccine-vaccinated adults with a 15 μg-BNT162b2 mRNA dose elicited high immune responses against the SARS-CoV-2 delta variant [6, 7]. Our previous study demonstrated that individuals who received inactivated vaccines as their primary series achieved a high immune response against delta and omicron variants following receipt of a booster vaccination with the standard dose of BNT162b2 or mRNA-1273 [1]. However, it remains unclear whether a half-dose booster of an mRNA vaccine, especially a 50 μg-mRNA-1273 vaccination following the inactivated vaccine primary series, could induce adequate neutralization of the SARS-CoV-2 delta and omicron variants. Our study compared the reactogenicity and immunogenicity of reduced and standard doses of the BNT162b2 and mRNA-1273 vaccines in CoronaVac (CV)-vaccinated individuals.

## 2. Materials and methods

### 2.1. Study Design

This prospective cohort study enrolled healthy Thai adults ≥18 years of age without previous COVID-19 infection by medical interview at the time of enrollment. Adults who were immunized with the two-dose CV 6 months (± 1 month) after the second dose and consented to receive the 15 μg-BNT162b2 (n=59) or 50 μg-mRNA-1273 (n=51) reduced dose of the mRNA COVID-19 vaccine booster between November to December 2021 were included in the study. The comparison group included individuals who received the standard dose of either BNT162b2 (Pfizer-BioNTech Inc., NY) (n=54) or mRNA-1273 (Moderna Inc., Cambridge, MA) (n=58) for a previous study conducted between September and December 2021 [1]. The study protocol was approved by the Institutional Review Board (IRB) in the Faculty of Medicine at Chulalongkorn University (IRB690/64). This trial was included on the Thai Clinical Trials Registry (TCTR 20210910002). Written informed consent was obtained from participants prior to enrollment. The study was conducted according to the Declaration of Helsinki and the principle of Good Clinical Practice Guidelines (ICH-GCP). Blood samples were collected before vaccination (V1, day 0, baseline) and after receipt of the booster dose (V2, the second visit day 14 ± 7 and V3, the third visit, day 28 ± 7) (Supplementary Figure S1).

### 2.2. Vaccines

The BNT162b (Pfizer-BioNTech Inc., New York, NY) contains 15 μg (reduced) or 30 μg (standard) of mRNA encoding SARS-CoV2 spike (S) protein [8]. The mRNA-1273 (Moderna Inc., Cambridge, MA) contains 50 μg (reduced) or 100 μg (standard) of mRNA encoding SARS-CoV2 S protein [9].

### 2.3. Reactogenicity Assessment

The subjects were continuously monitored for local, systemic, and any adverse events (AEs) following immunization (AEFIs) occurring within 7 days using an online or paper-based self-assessment questionnaire. The data collection protocol was given to participants by trained investigators during the initial visit.

### 2.4. Laboratory Measurements

Serum samples were collected for analysis of total immunoglobulin (Ig) specific to the receptor-binding domain (RBD) of the SARS-CoV-2 spike protein, the IgG anti-RBD, the IgG anti nucleoprotein (anti-N), the neutralization assay against delta (B.1.617.2) and omicron (B.1.1.529/BA.1) variants using surrogate virus neutralization test (sVNT), and the 50% focus reduction neutralization test (FRNT_50_) as previously described [1, 10]. The seropositivity rate of sVNT showed > 30% inhibition, and the seropositivity rate of FRNT_50_ was > 20-serum dilution. The assay used to measure the interferon-gamma (IFN-γ) SARS-CoV-2 releasing T cell response was performed on a heparinized whole blood sample as previously described [10].

### 2.5. Statistical Analysis

The sample size was calculated using G*power software version 3.1.9.6 (based on conventional effect size = 0.25, given significance level (α) = 0.05, power (1-β) = 0.8, numerator degree of freedom = 3, and number of groups = 4). The graphical representation and statistical analyses were carried out using GraphPad Prism version 9.3.1. Categorical analyses of age and sex were performed using the Chi-square test. Adverse events were analyzed using the risk difference with a 95% confidence interval (CI). IgG-specific RBDs and FRNT_50_ were designated as geometric mean titers (GMT) with a 95% CI. Other parameters were presented as medians with interquartile ranges. The differences in antibody titers, S/C, and percentage inhibition and IU/mL minus nil between groups were calculated using the Kruskal–Wallis or Wilcoxon signed-rank test (non-parametric) with multiple comparison adjustments. A *p*-value of <0.05 was considered statistically significant.

## 3. Results

### 3.1. Demographic data

A total of 222 healthy participants were analyzed, including 110 participants enrolled to receive the reduced vaccine dose and 112 participants who received the standard vaccine dose and were enrolled in a prior study in September 2021 [1]. The baseline demographic characteristics of participants in all four groups who received reduced or standard doses of the BNT162b2 or mRNA-1273 vaccines were generally comparable (Table 1). The mean age of participants who received the 15 μg-BNT162b2, 30 μg-BNT162b2, 50 μg-mRNA-1273, or 100 μg-mRNA-1273 vaccines were 39.1 ± 10.0, 41.6 ± 10.1, 38.7 ± 11.1, and 37.0 ± 10.5 years, respectively. No significant differences in age and sex were observed between the groups. Underlying diseases of the enrolled participants included allergy, diabetes mellitus, dyslipidemia, hypertension, thyroid, and other inactive diseases (Table 1). The median days to follow-up for the second visit were 18.55 (14–21), 14.3 (13–21), 15.5 (14–21), and 16 (14–20) days for the 15 μg-BNT162b2, 30 μg-BNT162b2, 50 μg-mRNA-1273, and 100 μg-mRNA-1273, respectively. The median days to follow-up for the second visit were 28.6 (27–30), 28.3 (27–35), 28.1 (28– 29), and 27.9 (22–35) days for the 15 μg-BNT162b2, 30 μg-BNT162b2, 50 μg-mRNA-1273, and 100 μg-mRNA-1273, respectively.

**Table 1.** Demographics and characteristics of the vaccinated cohorts

|  | *BNT162b2*<br>(15 µg) | *BNT162b2*<br>(30 µg)<sup>y</sup> | *mRNA-1273*<br>(50 µg) | *mRNA-1273*<br>(100 µg)<sup>y</sup> |
| --- | --- | --- | --- | --- |
| **Total number (n)** | 59 | 54 | 51 | 58 |
| **Mean age ± SD (years)** | 39.1±10.0 | 41.6±10.1 | 38.7±11.1 | 37.0±10.5 |
| **Sex** |  |  |  |  |
| **Male (%)** | 28 (47.5%) | 32 (59.3%) | 22 (43.1%) | 28 (47.8%) |
| **Female (%)** | 31 (52.5%) | 22 (40.7%) | 29 (56.8%) | 30 (52.2%) |
| **Underlying disease (%)** |  |  |  |  |
| **Allergy** | 4 (6.8%) | 4 (7.4%) | 5 (9.8%) | 4 (6.9%) |
| **Diabetes Mellitus** | 3 (5.1%) | 2 (3.7%) | - | 2 (3.4%) |
| **Dyslipidemia** | 3 (5.1%) | 4 (7.4%) | - | 1 (1.7%) |
| **Hypertension** | 5 (8.5%) | 6 (11.1%) | 3 (5.8%) | 4 (6.9%) |
| **Thyroid** | 1 (1.7%) | 2 (3.7%) | 1 (1.9%) | 1 (1.7%) |
| **Others ‡** | 2 (3.4%) | 3 (5.6%) | 3 (5.8%) | 2 (3.4%) |
| **Follow-up** |  |  |  |  |
| **Second visit (two weeks)** |  |  |  |  |
| **Mean (range) days** | 18.5 (14-21) | 14.3 (13-21) | 15.5 (14-21) | 16 (14-20) |
| **Lost to follow-up (n)** | 3 | 1 | 1 | 0 |
| **Third visit (four weeks)** |  |  |  |  |
| **Mean (range) days** | 28.6 (27-30) | 28.3 (27-35) | 28.1 (28-29) | 27.9 (22-35) |
| **Lost to follow-up (n)** | 3 | 0 | 5 | 2 |

‡ Inactive disease—no medication involving immunosuppressant. <sup>y</sup> Previous enrolled participants of standard dose vaccination [1].

### 3.2. Reactogenicity data

The most common solicited AE for all groups was injection site pain. Systemic events, including headache and myalgia. headache, joint pain, and diarrhea were more frequent in the 15 μg-than the 30 μg-BNT162b2 group, while joint pain and chilling were more frequent in the 100 μg-than the 50 μg-mRNA-1273 group (Figure 1 and Supplementary Figure S2). No severe AEs were reported.

**Figure 1.**
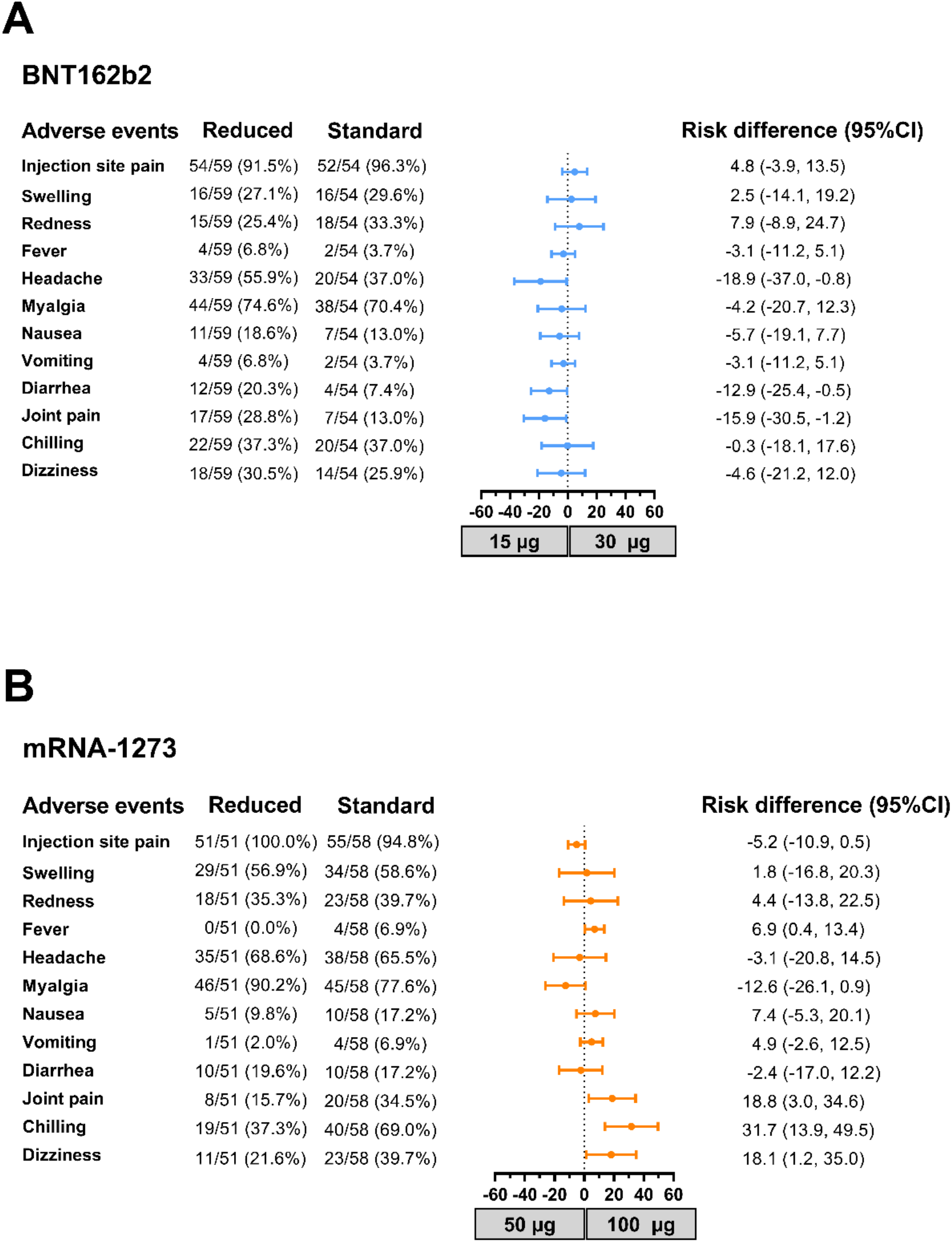
Reactogenicity data and risk difference analysis. The recorded incidence of AEs in participants who received the reduced booster dose with A) 15 μg (reduced) or 30 μg (standard) of BNT162b2 and B) 50 μg (reduced) or 100 μg (standard) of mRNA-1273. The risk difference with 95% CI is shown.

### 3.3. Ig Anti-N, total Ig, and IgG anti-RBD responses

Most participants were IgG anti-N seronegative (>94%) after receiving the mRNA vaccines (Supplementary Figure S3 and Table S1). There were no significant differences in the GMTs of total Ig and IgG anti-RBD between the four groups at baseline. By the second visit post-vaccination with 15 μg-BNT162b2, 30 μg-BNT162b2, 50 μg-mRNA-1273, and 100 μg-mRNA-1273, total Ig anti-RBD levels were 28,413, 31,793, 41,171, and 51,979 U/mL, respectively. By the third visit, total Ig anti-RBD was slightly reduced to 21,299, 21,053, 27,282, and 33,519 U/mL, respectively (Figure 2A). Moreover, the IgG anti-RBD levels were consistent with Ig anti-RBD (Figure 2B). These results indicated that there were no significant differences in Ig and IgG anti-RBD levels between reduced and standard doses of the same vaccine.

**Figure 2.**
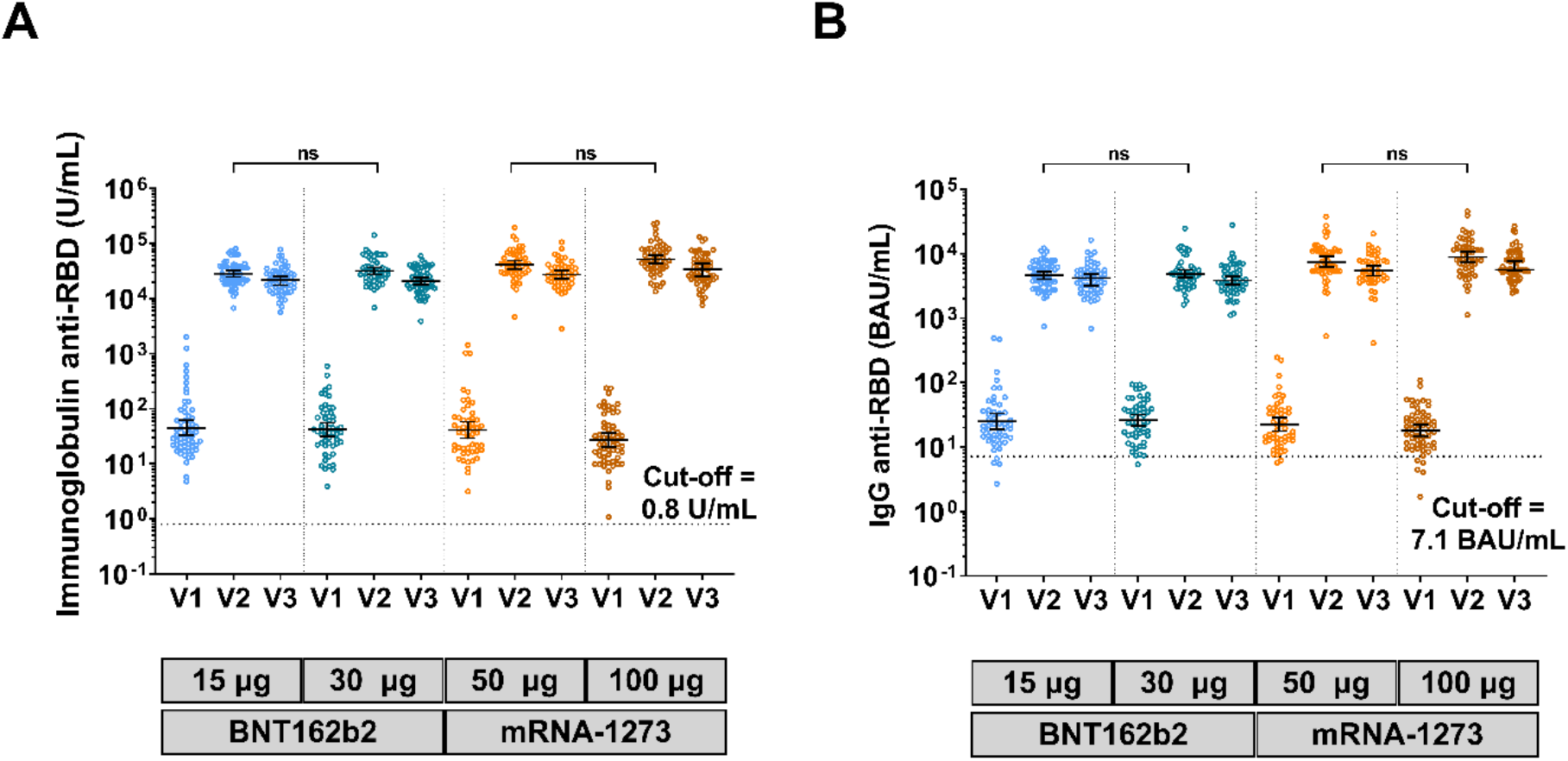
Antibody responses against SARS-CoV-2. A) Total immunoglobulin anti-RBD (U/mL) and B) IgG anti-RBD (BAU/mL). Serum was obtained from participants who received two completed doses of the inactivated vaccine, CoronaVac, followed by 15 μg (reduced) mRNA vaccine–BNT162b2, 30 μg (standard) mRNA vaccine–BNT162b2, 50 μg (reduced) mRNA-1273, or 100 μg (standard) mRNA-1273. Lines represent GMTs (95% CI); ns indicate no significant difference.

### 3.4. Neutralizing antibody against the SARS-CoV-2 delta and omicron variants

The sVNT against the delta and omicron variants was evaluated in a subgroup of participants. Seropositivity on the first visit was 3/40 (0.08%) and 1/40 (0.03%) to the delta and omicron variants, respectively. By the third visit, the sera of participants from all four groups achieved over 97% inhibition against the delta variant. In contrast, inhibition against the omicron variant at the third visit was 71.2%, 53.5%, 61.1%, and 76.7% in the 15 μg-BNT162b2, 30 μg-BNT162b2, 50 μg-mRNA-1273, and 100 μg-mRNA-1273 groups, respectively (Figure 3 and Supplementary Table S1). Inhibition against the omicron variant was significantly lower among participants in the 30 μg-BNT162b2 group than participants in the other groups (*p*<0.05) (Figure 3A).

**Figure 3.**
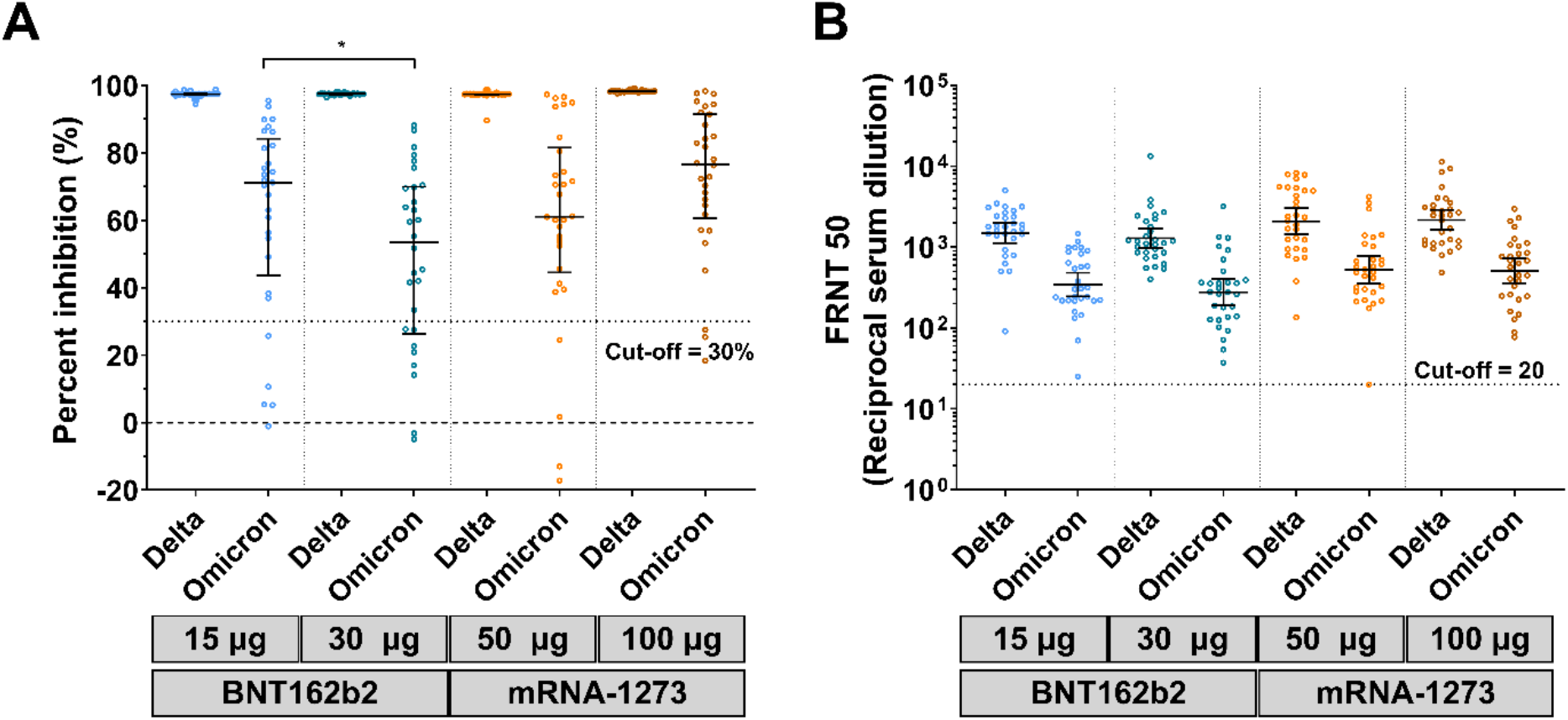
Neutralization assay against the SARS-CoV-2 delta (B.1.617.2) and omicron (B.1.1.529) variants. Serum samples were obtained from participants who received two completed doses of the inactivated vaccine, CoronaVac, followed by 15 μg (reduced) mRNA vaccine–BNT162b2, 30 μg (standard) mRNA vaccine–BNT162b2, 50 μg (reduced) mRNA-1273, or 100 μg (standard) mRNA-1273. A) Surrogate virus neutralization assay (sVNT); lines represent median (interquartile range). B) Focus reduction neutralization assay (FRNT_50_); lines represent GMTs (95% CI); ns indicates no significant difference; p<0.05 (*).

To assess live virus neutralization to the delta and omicron variants, FRNT_50_ was performed. On the first visit, most participants were seronegative for FRNT_50_ against the delta and omicron variants. By the third visit, the GMT against the delta variant in the 15 μg-BNT162b2 and 50 μg-mRNA-1273 groups increased to 1,505 and 2,088, respectively. The GMT against omicron in the 15 μg-BNT162b2 and 50 μg-mRNA-1273 groups was 343.4 and 541.2, respectively. Consistent with Ig RBD, no significant difference in FRNT_50_ was observed in sera from the reduced and standard-dose groups of the BNT162b2 and mRNA-1273 vaccines (Figure 3B and Supplementary Table S1).

### 3.5. QuantiFERON assay

The T-cell response was compared between the reduced and standard doses of the mRNA vaccines in participants who previously received the two-dose CoronaVac. Results showed that the median IFN-γ CD4+ T cell and CD4+ CD8+ T cell counts were higher at the second visit than at baseline in all groups. By the third visit, IFN-γ CD4+ and CD4+ CD8+ T cell counts were slightly reduced but this was not statistically significant. Importantly, there were no differences in T cell counts between participants receiving the reduced and standard doses of each vaccine (Figure 4A-B).

**Figure 4.**
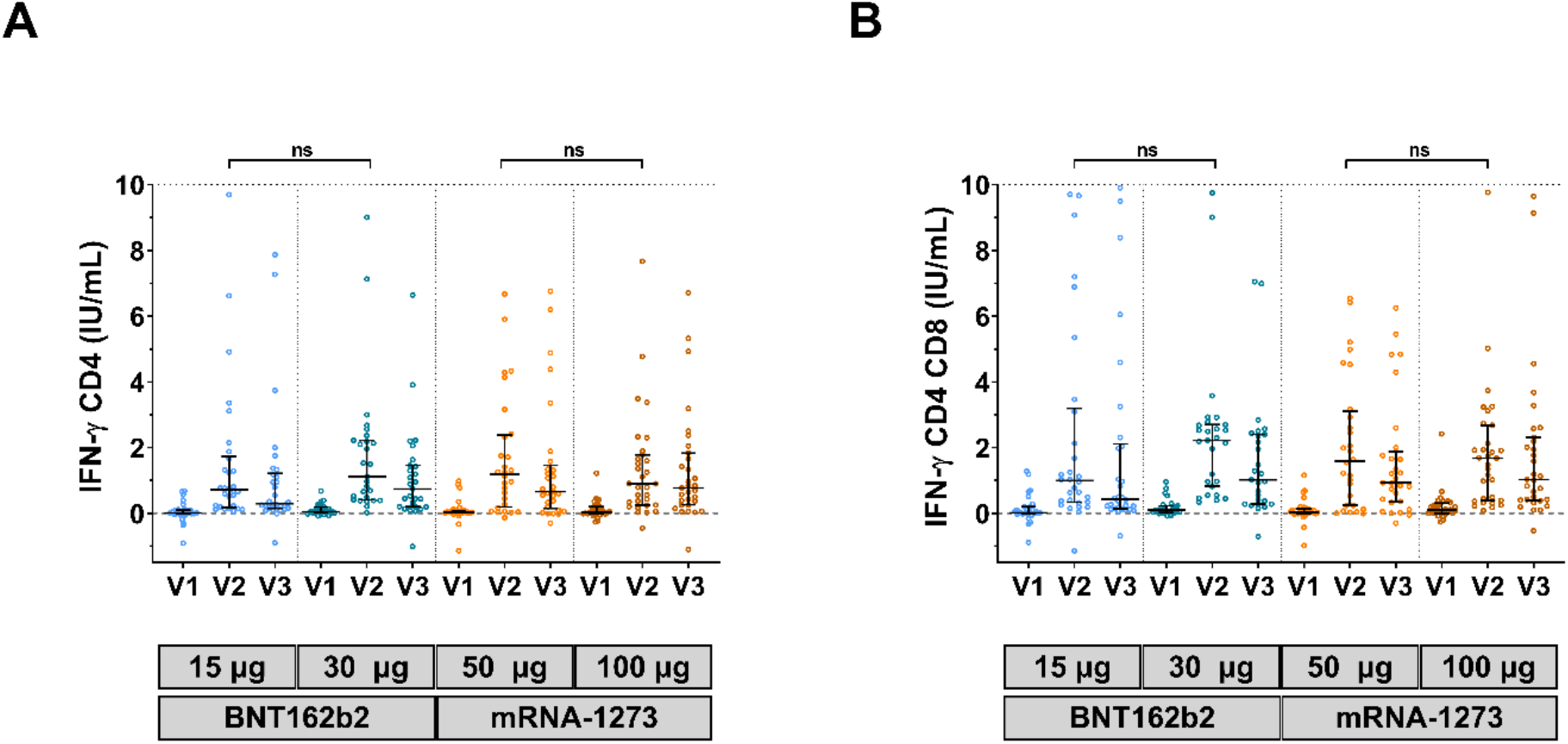
QuantiFERON SARS-CoV-2-stimulating interferon-gamma assay. Heparinized samples were immediately incubated in a QuantiFERON (QFN) blood collection tube for 21 h. The plasma was evaluated using QFN IFN-γ ELISA. A) IFN-γ was produced by CD4-specific Ag. B) IFN-γ was produced by CD4- and CD8-specific Ag2. Lines represent medians (IQR); ns indicate no significant difference.

## 4. Discussion and conclusion

This study compared the reactogenicity and immunogenicity of the reduced and standard doses of the mRNA vaccine booster in individuals who previously received a two-dose CV. A few participants had detectable IgG anti-N at baseline because they had received the two-dose CV. In agreement with Ig and IgG RBD responses, the reduced and standard-dose mRNA booster elicited high neutralizing activity against the SAR-CoV-2 delta and omicron variants. These results are consistent with a previous study of BNT162b2 following CV after three months [6]. Previous studies assessing the use of the standard mRNA vaccine dose after the CV series [1, 7] or three standard doses of BNT162b2 or mRNA-1273 [11] also demonstrated high neutralizing antibodies against the omicron variant. This aligns with another study reporting that the use of 15-μg-BNT162b2 as a booster dose elicited neutralizing activity against the SARs-CoV-2 delta and omicron variants [7]. Nevertheless, a previous study of mRNA-1273 showed that a third standard dose (100 μg) elicited a stronger immune response than a third reduced dose (50 μg) [12]. However, the reduced 50 μg-mRNA-1273 vaccine was able to elicit an immune response against the SARS-CoV-2 delta and omicron variants [12-13].

In the present study, IFN-γ release was observed by the second visit in participants who received the reduced mRNA vaccine dose. These results are consistent with previous studies of response to booster vaccination with the standard dose of mRNA vaccine after 3 [10] and 6 months [1] of CV priming. Both BNT162b2 and mRNA-1273 vaccines are able to stimulate robust antigen-specific T-cell responses after 7 days [14]. Indeed, BNT162b2 vaccination-induces T-cell responses, including induction of IFN-γ, TNF-α, and IL-2, and protection against ancestral strains might contribute to cross-protection against the omicron variant [15].

This study had some limitations. The surrogate virus neutralization test against the delta variant reached the upper limit of >97% inhibition. However, the FRNT_50_ result could explain why there were no significant differences in neutralizing activity against the delta and omicron variants. In addition, this study partially relied on subjects who received standard-dose immunization in another study. A more recent report of the standard-dose regimen should be used as the comparison group for the reduced-dose regimen. A follow-up study is underway to determine the durability of the booster vaccine.

In conclusion, there were no significant differences between administration of the reduced and standard doses of mRNA COVID-19 vaccine in participants who previously received inactivated CV vaccine. This study may inform decision-making regarding the use of reduced mRNA vaccine doses in healthy adults who were immunized with CV.

## Data Availability

The datasets generated and analyzed during the current study are available from the corresponding author on reasonable request.

## Acknowledgments

We would like to thank all Center of Excellence in Clinical Virology personnel and all participants for helping and supporting this project. This research was financially supported by the Health Systems Research Institute (HSRI), National Research Council of Thailand (NRCT), the Center of Excellence in Clinical Virology, Chulalongkorn University, and King Chulalongkorn Memorial Hospital, and partially supported by the Second Century Fund (C2F) of Sitthichai Kanokudom, Chulalongkorn University.

## Author Contributions

Conceptualization: S.K. (Sitthichai Kanokudom), P.N., S.H. and Y.P.; data collection:, D.S., R.Y. and T.T. (Thaksaporn Thatsanatorn); formal analysis: S.K. (Sitthichai Kanokudom); Investigation: S.K. (Sitthichai Kanokudom), N.S. (Natthinee Sudhinaraset), N.W. and Y.P.; methodology: S.K. (Sitthichai Kanokudom), S.A., N.S. (Nungruthai Suntronwong), J.C., C.A., T.T. (Thanunrat Thongmee), S.K. (Sirapa Klinfueng), T.D., W.C., P.P., P.V., and J.M.; project administration: Y.P.; writing—original draft: S.K.(Sitthichai Kanokudom), N.W., S.H. and Y.P.; writing—review and editing: S.K. (Sitthichai Kanokudom), N.W., S.H. and Y.P. All authors have read and agreed to the published version of the manuscript.

## Institutional Review Board Statement

The study protocol was approved by the Institutional Review Board (IRB), Faculty of Medicine, Chulalongkorn University (IRB number 690/64).

## Informed Consent Statement

Informed consent was obtained before participant enrollment. The study was conducted according to the Declaration of Helsinki and the Good Clinical Practice Guidelines (ICH-GCP) principles.

## Supplementary information

**Supplementary Figure S1**. Participant flow diagram. The cohorts were primed with the primary series of CV and received a reduced dose (experimental group) or standard dose (comparative group) of the mRNA vaccine.

**Supplementary Figure S2**. Reactogenicity after receiving mRNA vaccine for 7 days post-vaccination. The participants received two completed doses of the inactivated vaccine, CoronaVac, followed by A) 15 μg-BNT162b2, B) 30 μg-BNT162b2, C) 50 μg-mRNA-1273, or D) 100 μg-mRNA-1273. Participant percentages are shown on the Y-axis. Swelling and redness were graded by measuring the diameter area as mild (<5 cm), moderate (5 to <10 cm), and severe (≥10 cm). Fever was defined as mild (38.0-38.5 °C), moderate (38.5-39.0 °C), and severe (39.0 °C). Local and systemic symptoms were graded as mild (easily tolerated with no limitation on regular activity), moderate (some limitation of daily activity), and severe (unable to perform the regular daily activity).

**Supplementary Figure S3**. IgG anti nucleoprotein (N) of SARS-CoV-2 index (S/C). Serum samples were obtained from participants who received two completed doses of the inactivated vaccine, CoronaVac, followed by 15 μg (reduced) mRNA vaccine–BNT162b2, 30 μg (standard) mRNA vaccine–BNT162b2, 50 μg (reduced) mRNA-1273, or 100 μg (standard) mRNA-1273. Lines represent the median (interquartile range).

**Supplementary Table S1**. Patient serum responses during study trials.

