## Supplementary information for "Comparison of the reactogenicity and immunogenicity of a reduced and standard booster dose of the mRNA COVID-19 vaccine in healthy adults after two doses of inactivated vaccine"

**Supplementary Figure S1.** Participant flow diagram. The cohorts were primed with the primary series of CV and received a reduced dose (experimental group) or standard dose (comparative group) of the mRNA vaccine.

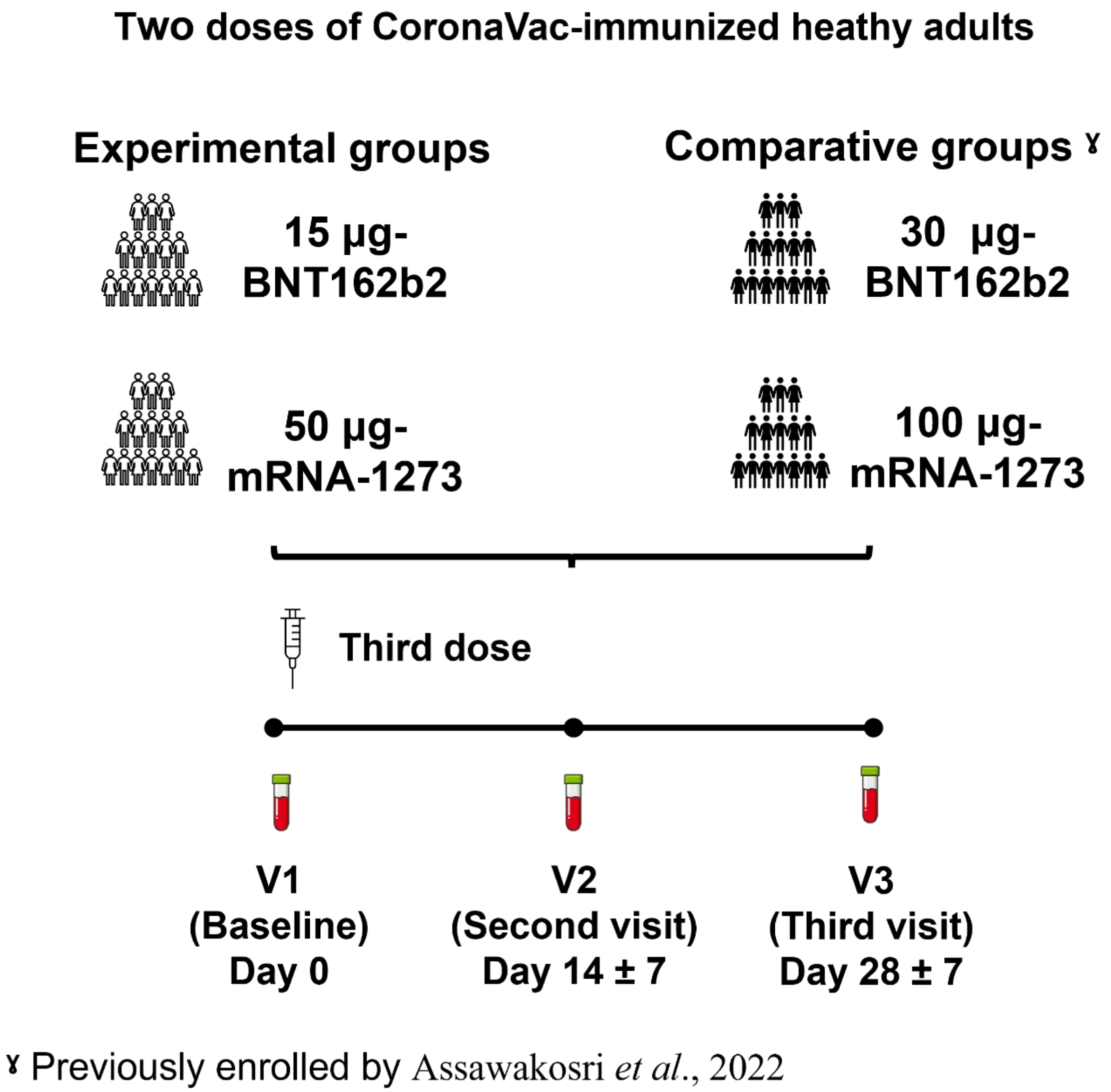

**Supplementary Figure S2.** Reactogenicity after receiving mRNA vaccine for 7 days post-vaccination. The participants received two completed doses of the inactivated vaccine, CoronaVac, followed by A) 15 μg-BNT162b2, B) 30 μg-BNT162b2, C) 50 μg-mRNA-1273, or D) 100 μg-mRNA-1273. Participant percentages are shown on the Y-axis. Swelling and redness were graded by measuring the diameter area as mild (<5 cm), moderate (5 to <10 cm), and severe (≥10 cm). Fever was defined as mild (38.0-38.5 ◦C), moderate (38.5-39.0 ◦C), and severe (39.0 ◦C). Local and systemic symptoms were graded as mild (easily tolerated with no limitation on regular activity), moderate (some limitation of daily activity), and severe (unable to perform the regular daily activity).

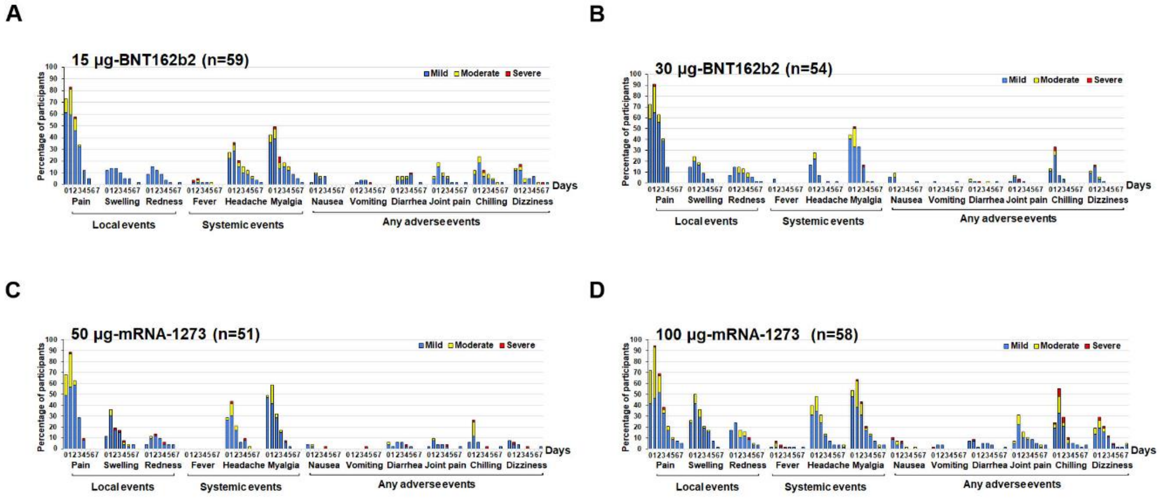

**Supplementary Figure S3.** IgG anti nucleoprotein (N) of SARS-CoV-2 index (S/C). Serum samples were obtained from participants who received two completed doses of the inactivated vaccine, CoronaVac, followed by 15 μg (reduced) mRNA vaccine–BNT162b2, 30 μg (standard) mRNA vaccine–BNT162b2, 50 μg (reduced) mRNA-1273, or 100 μg (standard) mRNA-1273. Lines represent the median (interquartile range).

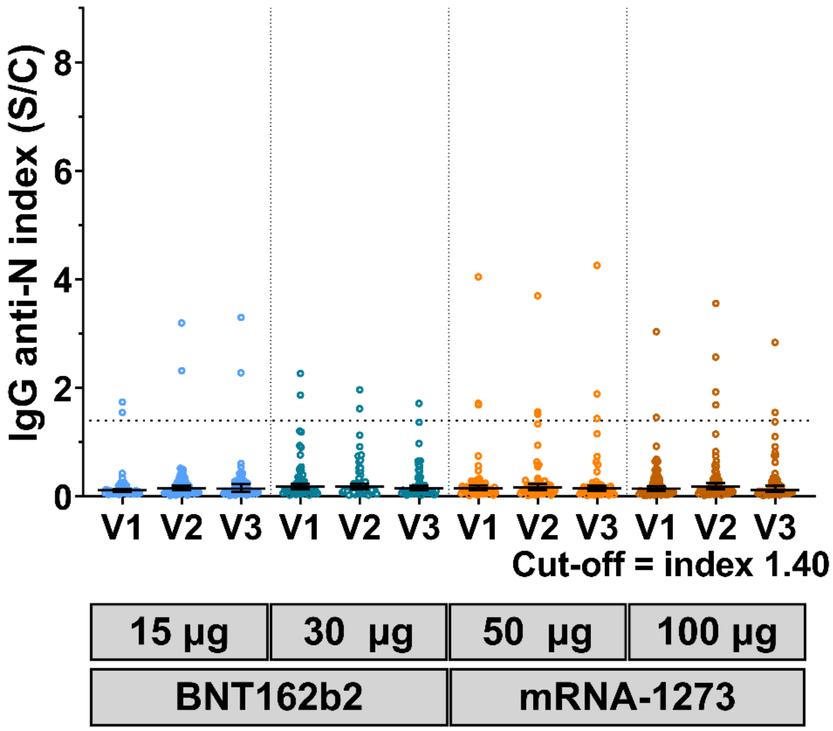

**Supplementary Table S1**. Patient serum responses during study trials.

| | BNT162b2 (15 µg) | BNT162b2 (30 µg)[¥] | mRNA-1273 (50 µg) | mRNA-1273 (100 µg)[¥] |
|---|---|---|---|---|
| **Humoral responses** | | | | |
| **Ig anti-RBD (U/mL)** | | | | |
| V1 (Baseline), n | 59 | 54 | 51 | 58 |
| GMT (95% CI) | 45.01 (32.86-61.65) | 41.67 (31.38-55.32) | 41.06 (29.05-58.03) | 27.07 (20.44-35.84) |
| V2 (Second visit), n | 56 | 54 | 51 | 57 |
| GMT (95% CI) | 28413 (24781-32578) | 31793 (27428-36852) | 41171 (34563-49043) | 51979 (43920- 61516) |
| V3 (Third visit), n | 56 | 54 | 49 | 56 |
| GMT (95% CI) | 21299 (18390-24667) | 21053 (18145-24428 | 27282 (23082-32245) | 33519 (28228- 39802) |
| **IgG anti-RBD (BAU/mL)** | | | | |
| V1 (Baseline), n | 59 | 54 | 51 | 58 |
| GMT (95% CI) | 22.91 (17.88-29.37) | 26.01 (21.16-31.97) | 22.52 (17.70-28.64) | 18.13 (14.78-22.23) |
| V2 (Second visit), n | 56 | 54 | 51 | 57 |
| GMT (95% CI) | 4609 (4009-5299) | 4878 (4222-5636) | 7476 (6167-9062) | 8901 (7471-10605) |
| V3 (Third visit), n | 56 | 54 | 49 | 56 |
| GMT (95% CI) | 3994 (3455-4618) | 3837 (3292-4472) | 5416 (4523-6485) | 6570 (5694-7580) |
| **IgG anti-N (S/C)** | | | | |
| V1 (Baseline), n | 59 | 54 | 51 | 58 |
| Median (IQR) | 0.09 (0.07-0.19) | 0.15 (0.10-0.36) | 0.15 (0.09-0.23) | 0.13 (0.07-0.33) |
| V2 (Second visit), n | 56 | 54 | 51 | 57 |
| Median (IQR) | 0.15 (0.08-0.29) | 0.14 (0.09-0.41) | 0.17 (0.09-0.25) | 0.14 (0.09-0.37) |
| V3 (Third visit), n | 56 | 54 | 49 | 56 |
| Median (IQR) | 0.15 (0.07-0.27) | 0.12 (0.08-0.31) | 0.15 (0.07-0.22) | 0.12 (0.07-0.31) |
| **Neutralization assays: VNT-Delta (%inhibition)** | | | | |
| V3 (Third visit), n | 29 | 30 | 30 | 30 |
| Median (IQR) | 97.4 (97.2-97.5) | 97.5 (97.3-97.6) | 97.4 (97.2-97.4) | 98.2 (98.2-98.3) |
| **Neutralization assays: VNT-Omicron (%inhibition)** | | | | |
| V3 (Third visit), n | 29 | 30 | 30 | 30 |
| Median (IQR) | 71.2 (43.7-84.2) | 53.5 (26.3-70.0) | 61.1 (44.5-81.5) | 76.7 (60.6-91.6) |
| **Neutralization assay: FRNT-Delta (dilution)** | | | | |
| V3 (Third visit), n | 30 | 30 | 30 | 30 |
| GMT (95% CI) | 1505 (1126- 2012) | 1285 (982.2-1680) | 2088 (1441-3025) | 2168 (1627-2889) |
| **Neutralization assay: FRNT-Omicron (dilution)** | | | | |
| V3 (Third visit), n | 30 | 30 | 30 | 30 |
| GMT (95% CI) | 343.4 (244.6- 482.1) | 276.6 (190.1-402.3) | 521.1 (352.4-770.7) | 512.5 (358.7-732.2) |

| | | | | |
|---|---|---|---|---|
| **T-cell responses** | | | | |
| **IFN-γ CD4⁺ (IU/mL)** | | | | |
| V1 (Baseline), n | 30 | 27 | 29 | 31 |
| Median (IQR) | 0.01 (-0.02-0.103) | 0.05 (0.01-0.19) | 0.03 (0.01-0.08) | 0.03 (-0.01-0.20) |
| V2 (Second visit), n | 30 | 27 | 28 | 31 |
| Median (IQR) | 0.72 (0.18 - 1.72) | 1.12 (0.41-2.22) | 1.18 (0.19-2.39) | 0.90 (0.25-1.78) |
| V3 (Third visit), n | 30 | 27 | 29 | 31 |
| Median (IQR) | 0.30 (0.15-1.22) | 0.74 (0.20-1.46) | 0.66 (0.15-1.46) | 0.77 (0.26-1.83) |
| **IFN-γ $CD4^+$ $CD8^+$ (IU/mL)** | | | | |
| V1 (Baseline), n | 30 | 27 | 29 | 31 |
| Median (IQR) | 0.02 (-0.02-0.20) | 0.10 (0.02-0.23) | 0.03 (-0.01-0.13) | 0.11 (0.00-0.31) |
| V2 (Second visit), n | 30 | 27 | 28 | 31 |
| Median (IQR) | 1.00 (0.34-3.19) | 2.22 (0.82-2.70) | 1.59 (0.24-3.11) | 1.68 (0.39-2.68) |
| V3 (Third visit), n | 30 | 27 | 29 | 31 |
| Median (IQR) | 0.44 (0.14-2.11) | 1.02 (0.29-2.40) | 0.93 (0.35-1.88) | 1.03 (0.40-2.31) |

µg indicates an amount of SARS-CoV-2 Spike protein encoded mRNA

[ɣ] the comparative group represents a regular dose of each mRNA COVID-19 vaccine as previously reported by Assawakosri et al., 2011.
